# Distinct Mucoinflammatory Phenotype and the Immunomodulatory Long Noncoding Transcripts Associated with SARS-CoV-2 Airway Infection

**DOI:** 10.1101/2021.05.13.21257152

**Authors:** Dinesh Devadoss, Arpan Acharya, Marko Manevski, Kabita Pandey, Glen M. Borchert, Madhavan Nair, Mehdi Mirsaeidi, Siddappa N. Byrareddy, Hitendra S. Chand

**Affiliations:** Department of Immunology and Nano-Medicine, Herbert Wertheim College of Medicine, Florida International University, Miami, FL-33199; Department of Pharmacology and Experimental Neuroscience, University of Nebraska Medical Center, Omaha, NE-68198; Department of Pharmacology, University of South Alabama, Mobile, AL-36688; Miller School of Medicine, Division of Pulmonary, Critical Care, and Sleep Medicine, University of Miami, Miami, FL-33136

**Author notes:** **Address for Correspondence:** Hitendra S. Chand, PhD, Department of Immunology and Nano-Medicine, Herbert Wertheim College of Medicine, Florida International University, Miami, FL 33199, USA. **Funding Support:** NIH AI144374, AI152937, and DA052845.

**Keywords:** Respiratory epithelial cells, SARS-CoV-2, COVID-19, long noncoding RNAs, mucoinflammatory response

## Abstract

Respiratory epithelial cells are the primary target for severe acute respiratory syndrome coronavirus 2 (SARS-CoV-2). We investigated the 3D human airway tissue model to evaluate innate epithelial cell responses to SARS-CoV-2 infection. A SARS-CoV-2 clinical isolate productively infected the 3D-airway model with a time-dependent increase in viral load (VL) and concurrent upregulation of airway immunomodulatory factors (*IL-6, ICAM-1*, and *SCGB1A1*) and respiratory mucins (*MUC5AC, MUC5B, MUC2*, and *MUC4)*, and differential modulation of select long noncoding RNAs (lncRNAs i.e., *LASI, TOSL, NEAT1*, and *MALAT1*). Next, we examined these immunomodulators in the COVID-19 patient nasopharyngeal swab samples collected from subjects with high- or low-VLs (∼100-fold difference). As compared to low-VL, high-VL patients had prominent mucoinflammatory signature with elevated expression of *IL-6, ICAM-1, SCGB1A1, SPDEF, MUC5AC, MUC5B*, and *MUC4*. Interestingly, *LASI, TOSL*, and *NEAT1* lncRNA expressions were also markedly elevated in high-VL patients with no change in *MALAT1* expression. In addition, dual-staining of *LASI* and SARS-CoV-2 nucleocapsid *N1* RNA showed predominantly nuclear/perinuclear localization at 24 hpi in 3D-airway model as well as in high-VL COVID-19 patient nasopharyngeal cells, which exhibited high MUC5AC immunopositivity. Collectively, these findings suggest SARS-CoV-2 induced lncRNAs may play a role in acute mucoinflammatory response observed in symptomatic COVID-19 patients.

## Introduction

Initial host-viral interactions in the upper respiratory tract critically affect subsequent respiratory and systemic immune responses to other co-infections or environmental exposures. SARS-CoV-2 gains entry via the respiratory mucosa and any host factor dysregulations occurring during this interaction can result in pulmonary and/or extrapulmonary complications (1, 2). Notably, the proteins necessary for viral entry are highly abundant in respiratory epithelial cells (RECs) (1, 3, 4), rendering the respiratory tract highly susceptible to SARS-CoV-2 infection. SARS-CoV-2 is an enveloped β-coronavirus with a heavily glycosylated trimer spike (S) protein which facilitates viral attachment to ACE2 (angiotensin-converting enzyme 2) or CD147 (Basigin or BSG) (5, 6). Following attachment, trimming of S protein by host cellular proteases such as TMPRSS2 (transmembrane serine protease 2) and furin mediates viral entry (7-9). That said, the expression of these viral receptors is sensitive to various immunomodulators. For example, in addition to being the protective barrier and a competent airway lumen clearance mechanism, airway mucins normally moderate the mucosal immune responses (10). During SARS-CoV-2 infection, however, induced inflammatory factors drive airway tissue remodeling and severe inflammation can cause mucus hyperexpression potentially leading to acute respiratory distress syndrome (ARDS) (11). Managing this immune response constitutes of the major challenges faced in effectively treating COVID-19 for patients presenting with severe manifestations.

Analyzing host-viral interactions is vital for understanding viral pathogenesis, and while several notable protein interactions necessary for SARS-CoV-2 infection and/or progression are now well documented, documented roles for specific host long noncoding RNAs (lncRNAs) during SARS-CoV-2 infection remain elusive. LncRNAs regulate virtually every cellular function including moderating the immune response of infected host cells and regulating the viral genome packing and replication (12, 13). Although a few studies have reported on the expression profiling of lncRNAs in stratifying the COVID-19 disease severity and associated immune dysfunction (14-17), there are limited studies on the respiratory epithelia specific lncRNAs and their role in SARS-CoV-2 infection (18, 19). As such, in this study, we assessed acute mucoinflammatory and immunomodulatory responses, including lncRNA expressions in a 3D airway tissue model of acute SARS-CoV-2 infection and COVID-19 patient nasopharyngeal samples to gain novel insights into early innate responses of human respiratory cells.

## Results and Discussion

### SARS-CoV-2 productively infects a 3D airway tissue model and induces inflammatory and mucous responses

To model the acute respiratory infection conditions, a 3D airway tissue culture model of primary human respiratory tract epithelial cells (RTECs) was infected with SARS-CoV-2 primary isolate (USA-WI1/2020, BEI Cat # NR-52384) at 1 multiplicity of infection (MOI) (see Supplemental Methods for details). Viral RNA was assessed in apical washes, in the basal culture supernatant, and in cells; from RNA samples extracted at 0, 1, 4, 24, and 48 h post-infection (hpi). Viral RNA increased by 3.5-fold from 8.9×10^5^ viral genomic equivalents per ml at 1 hpi to 3.1×10^6^ at 24 hpi in apical washes (Figure 1A), there was no change in VL in basal culture supernatant which remained at ∼3.0×10^5^ genomic equivalents per ml at all the time-points (Figure 1B), and the level of vRNA in total cell RNA increased by ∼6 fold at 24 hpi (Figure 1C). Thus, 3D airway cells productively infected with an actively replicating SARS-CoV-2 shed the virions almost exclusively from the apical surface.

**Figure 1.**
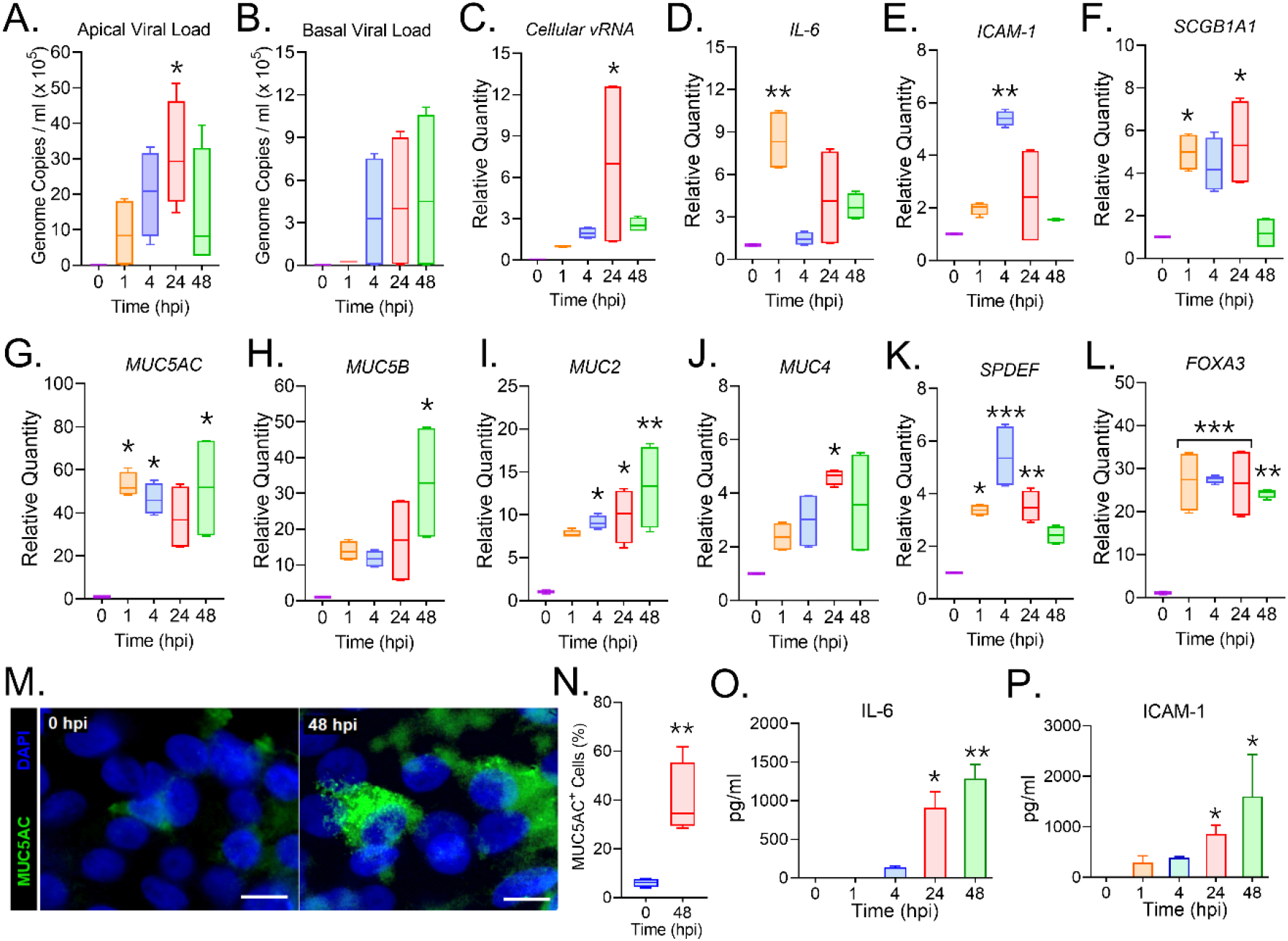
SARS-CoV-2 infection of human respiratory epithelial cells induces a robust mucoinflammatory phenotype in a 3D airway tissue model. Respiratory tract epithelial cells (RTECs) differentiated on air-liquid interface were infected with 1 MOI of SARS-CoV-2 clinical isolate (USA-WA1/2020 isolate) and analyzed at 0, 1, 4, 24, and 48 h post-infection (hpi). Viral loads determined in **(A.)** the apical wash, **(B.)** the basal culture supernatant, and **(C.)** in the cell RNA. Relative expression levels of inflammatory factors **(D.)** *IL-6* and **(E.)** *ICAM-1* mRNAs, and **(F.)** the epithelial secretory factor *SCGB1A1* mRNA. Changes in the mRNA expression of airway mucins, **(G.)** *MUC5AC*, **(H.)** *MUC5B*, **(I.)** *MUC2*, and **(J.)** *MUC4* mRNA levels; and the mucin regulatory transcriptional factors, **(K.)** *SPDEF and* **(L.)** *FOXA3* as analyzed by qPCR of the total cell RNA. **(M.)** Representative micrographs of RTECs at 0 and 48 hpi showing MUC5AC (shown in green) immunoreactivity along with the DAPI stained nuclei (shown in blue), scale - 5µ. **(N.)** Percentage of MUC5AC+ cells within each treatment group. Quantification of **(O.)** IL-6 and **(P.)** ICAM-1 levels in media supernatants as assessed by ELISA (n=4/gp from 2 independent experiments; *p<0.05; \*\**p*<0.01; \*\*\**p*<0.001 by ANOVA).

In addition, we found the expression of the viral entry host factor, *ACE2*, significantly reduced at all time points following infection (see Supplemental Figure 1A). In contrast, the expression of S protein processing enzyme *TMPRSS2* remained constant for first 24 hpi but was markedly suppressed at 48 hpi (Supplemental Figure 1B). Thus, SARS-CoV-2 infection directs ACE2 and TMRPSS2 suppression in a RTEC 3D model, as reported earlier (8, 20). We also found altered expressions of several inflammatory factors charged with directing airway tissue remodeling and/or controlling host immune responses and airway mucin expression (21, 22). Among the inflammatory factors, although CXCL-8 expression was unaffected following infection (Supplemental Figure 1C), interleukin-6 (*IL-6)* was significantly elevated (9-fold) at 1 hpi (Figure 1D), whereas intercellular adhesion molecule-1 (*ICAM-1)* was elevated (>5-fold) at 24 hpi (Figure 1E) and airway secretoglobulin 1A1 (*SCGB1A1)*, also known as club cell secretory protein (CCSP) was induced by 5-fold from 1 to 24 hpi with then returned to baseline levels at 48 hpi (Figure 1F).

Notably, we also observed robust mucin hyperexpression in our 3D airway tissue model. SARS-CoV-2 infection robustly upregulated secretory mucin *MUC5AC* (21, 23-26) transcription by more than 50-fold by 1 hpi and maintained hyperexpression at all the time-points (Figure 1G). In addition, expression of the constitutive epithelial mucin, *MUC5B*, involved in the airway antimicrobial response (27-29) was increased by 48 hpi (Figure 1H), and similar to *MUC5AC*, cell surface-associated mucins, *MUC2* (Figure 1I), and *MUC4* (Figure 1J) levels were also increased by 1 hpi then remain elevated at all time-points. Furthermore, known transcriptional regulators of mucin expression (30, 31) were similarly upregulated following infection with >2-fold induction of *SPDEF* levels (Figure 1K) and >20-fold induction of *FOXA3* levels at 1 hpi (Figure 1L).

Next, protein expressions were assessed in transwell cells by immunostaining. In agreement with our quantitative RT-PCR analyses, immunostaining confirmed robust immunopositivity for MUC5AC expression (Figure 1M) with ∼40% MUCAC+ cells at 48 hpi compared to <5% cells at 0 hpi (Figure 1N). Levels of secreted inflammatory factors, IL-6 (Figure 1O) and ICAM-1 (Figure 1P) in basal media supernatants were also elevated at 24 and 48 hpi. Thus, the inflammatory and mucous responses of respiratory epithelial cells are acutely moderated by SARS-CoV-2 infection. Although roles for airway mucins in innate antiviral immunity are well documented, SARS-CoV-2 associated mucin signatures have not been thoroughly evaluated to date (32). Specifically, these mucins are heavily glycosylated and could result in higher viral binding/retention due to interaction with sialic acids (33) and contribute to viral infection and/or transmission.

### SARS-CoV-2 infected airway tissue model show increased expression of immunomodulatory lncRNAs

In addition to mucoinflammatory factor encoding mRNAs, another type of RNA, lncRNAs, have now been found to play vital roles in regulating the innate immune responses (12, 13) and recent studies have shown that specific lncRNAs are differentially expressed during host-pathogen interactions following viral infection (18, 19, 34, 35). That said, our group identified two novel epithelial lncRNAs, *LASI* (lncRNA on antisense strand to ICAM-1) and *TOSL* (TNFAIP3-opposite strand lncRNA) that are differentially expressed during the mucus hypersecretory response (36) and blocking these lncRNA expression alleviates the mucin hyperexpression. Notably, we find *LASI* transcripts are highly elevated (10-fold) at 1 hpi (Figure 2A), whereas *TOSL* expression is increased by 1 hpi and were >6-fold higher at 48 hpi (Figure 2B). However, in contrast to recent clinical reports (19, 37, 38), we find NEAT1 (Nuclear Enriched Abundant Transcript 1) lncRNA levels reduced following infection (Figure 2C), and no change in the expression of MALAT1 (Metastasis Associated Lung Adenocarcinoma Transcript 1) another immunomodulatory lncRNA (39, 40) MALAT1 (Metastasis Associated Lung Adenocarcinoma Transcript 1) expression (Supplemental Figure 1D). In agreement with our qRT-PCR based expression, dual-RNA FISH analysis showed co-expression of SARS-CoV-2 *N1 vRNA* and *LASI* transcripts around the nuclear/perinuclear region (Figure 2D) with elevated expressions at 24 hpi compared to 4 hpi as assessed by the histological-score (H-score) quantitation H-score (Figure 2E). Thus, SARS-CoV-2 infection of our 3D airway culture model resulted in a strong mucoinflammatory phenotype with concurrent increases in associated lncRNA expressions. Therefore, our data suggest that these lncRNAs may be serve as biomarkers for measuring SARS-CoV-2 disease progression. However, these need to be confirmed using large number of clinical samples.

**Figure 2.**
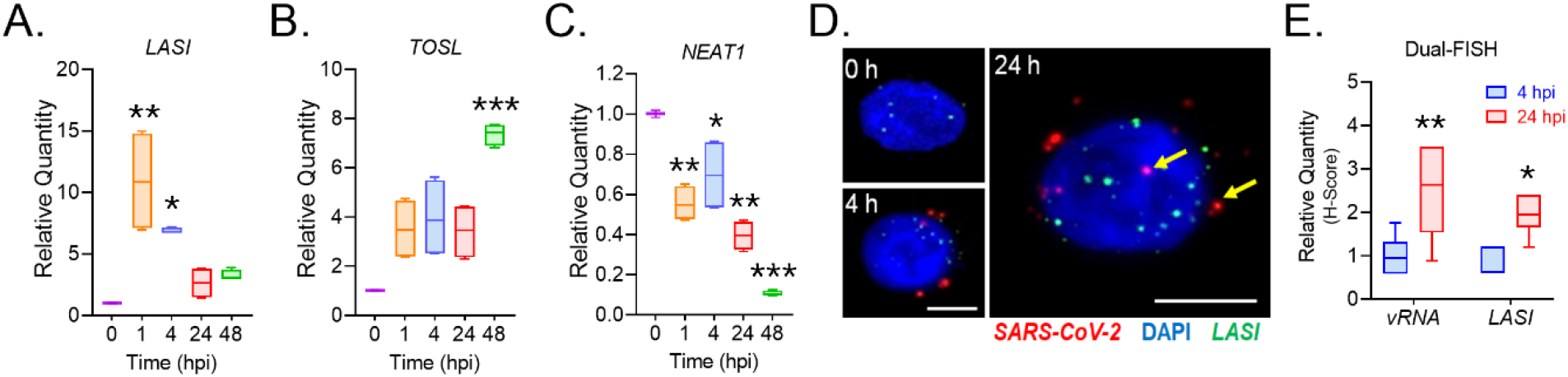
SARS-CoV-2 infection differentially affects the expression of inflammation associated lncRNAs in airway epithelial cells. Relative expression levels of select immunomodulatory lncRNAs, **(A.)** *LASI*, **(B.)** *TOSL*, and **(C.)** *NEAT1* in airway epithelial cells following 0, 1, 4, 24, and 48 h of SARS-CoV-2 infection. (n=4/gp from 2 independent experiments; *p<0.05; \*\**p*<0.01; \*\*\**p*<0.001 by ANOVA). **(D.)** Colocalization of SARS-CoV-2 *vRNA* and *LASI* transcripts in infected cells at 0, 4, and 24 hpi as determined by the dual-FISH analysis and structured illumination microscopy. Representative micrograph of dual-FISH-stained cells showing SARS-CoV-2 nucleocapsid *vRNA* (in red) and *LASI* transcripts (in green) along with DAPI-stained nuclei (in blue), scale – 2 µ. **(E.)** H-score quantitation of *vRNA* and *LASI* transcripts per cell at 4 and 24 hpi (n=9-10 cells/gp; *p<0.05; \*\**p*<0.01; by Student’s t-test).

### COVID-19 patients with higher viral load show increased mucoinflammatory response

Large cohort studies revealed there was no difference in SARS-CoV-2 viral load between symptomatic and asymptomatic patients, whereas disease severity and mortality in symptomatic patients is directly correlated with viral load (38, 41, 42). The mucosal immune response of upper respiratory tract plays a critical role in viral shedding and replication. To explore this correlation, in our study, we divided subjects into 2 groups based on the qRT-PCR cycle threshold (C_T_) of SARS-CoV-2 N1 nucleocapsid, with a C_T_>30 considered a low viral load (Lo-VL) and a C_T_<30 high viral load (Hi-VL). As detailed in Supplemental Table 1, nasopharyngeal swabs collected from a total of 20 subjects (10 Hi-VL and 10 Lo-VL) were examined. Twelve symptomatic subjects were hospitalized and had at least one and/or combination of comorbidities *viz*., obesity, chronic disorders, hypertension, and diabetes. Hi-VL subjects were found to contain >100-fold higher *vRNA* levels than Lo-VL subjects (Figure 3A). Interestingly, no significant differences were observed in *ACE2* (Figure 3B) and *TMPRSS2* (Figure 3C) mRNA levels between groups. In contrast, Hi-VL samples exhibited ∼1.5-fold higher *IL-6* (Figure 3D) and *ICAM-1* (Figure 3E) inflammatory factors mRNA expressions, as well as 4-fold higher *SCGB1A1* expression (Figure 3F). Hi-VL patient samples also exhibited robust secretory mucin levels, with ∼10-fold higher *MUC5AC* (Figure 3G) and 4-fold higher *MUC5B* (Figure 3H) expressions. Notably, there was a trend towards reduced MUC2 expression (Figure 3I) in Hi-VL subjects with ∼10-fold higher *MUC4* (Figure 3J) expression compared to Lo-VL subjects. Expression of *SPDEF* was also increased by ∼5-fold in Hi-VL subjects (Figure 3K), but in contrast to our 3D models, we failed to detect any FOXA3 transcripts in patient nasopharyngeal swab samples. These data clearly suggests that SARS-CoV-2 can regulate the expression of several cellular receptors by inducing inflammatory responses.

**Figure 3.**
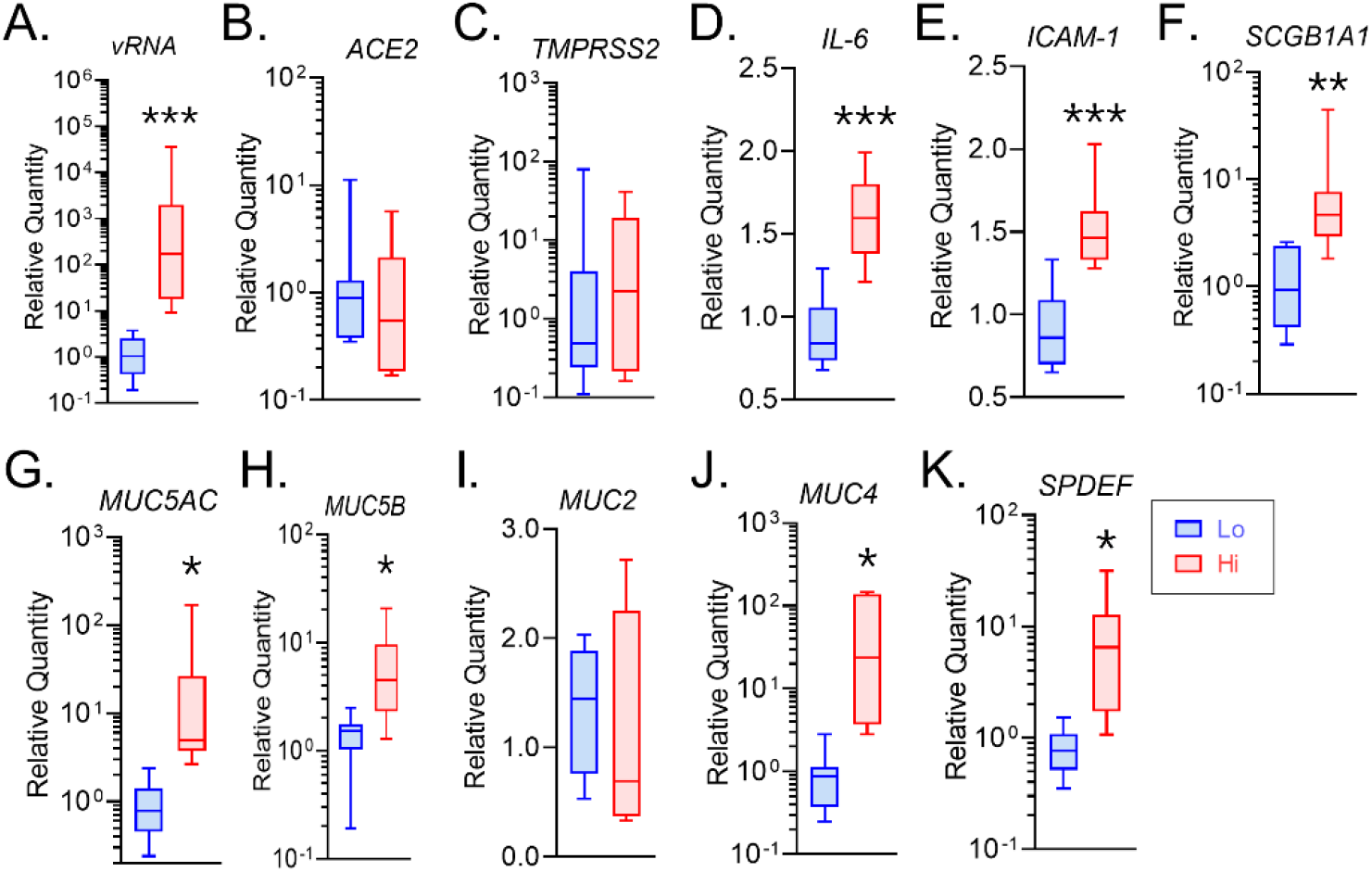
COVID-19 patients with high nasopharyngeal viral load show increased mucoinflammatory phenotype compared to the patients with low viral load. Total RNA from nasopharyngeal swab samples of COVID-19 patients were analyzed. Based on the SARS-CoV-2 nucleocapsid viral RNA expression, the patients with high viral load (Hi-VL) had average C_T_ values of 25.9±1.3 whereas those with low viral load (Lo-VL) had average C_T_ values of 35.5±0.7 (n=10/gp). Relative mRNA expression of: **(A.)** SARS-CoV-2 viral RNA (*vRNA*) and viral entry related host factors, **(B.)** *ACE2* and **(C.)** *TMPRSS2* mRNA; Innate inflammatory factors, **(D.)** *IL-6*, **(E.)** *ICAM-1*, and **(F.)** *SCGB1A1*; Airway mucins **(G.)** *MUC5AC*, **(H.)** *MUC5B*, **(I.)** *MUC2*, and **(J.)** *MUC4;* and **(K.)** *SPDEF* transcriptional factor in the Hi-VL vs Lo-VL patient swab samples (n=10/gp; *p<0.05; \*\**p*<0.01; \*\*\**p*<0.001 by Student’s t-test).

### Respiratory mucosa lncRNAs are associated with SARS-CoV-2 viral load

While we found no difference in MALAT1 lncRNA levels between Hi-VL and Lo-VL subjects (Supplemental Figure 2A), the expression of select immunomodulatory lncRNAs, *LASI* (Figure 4A), *TOSL* (Figure 4B), and *NEAT1* (Figure 4C) were significantly upregulated in Hi-VL patient samples. In addition, dual-RNA FISH analysis of nasal swab epithelial cells from Hi-VL subjects showed enriched SARS-CoV-2 nucleocapsid *vRNA* and *LASI* transcripts in nuclear and cytosolic regions (Figure 4D). The H-score quantitation confirmed *vRNA* and *LASI* transcripts were 1.5-fold higher in Hi-VL samples compared to Lo-VL samples (Figure 4F), corroborating our qRT-PCR data. Nasal epithelial cell mucin protein expression was also analyzed for the mucin protein expression by immunostaining for MUC5AC (Figure 4E) and MUC5B (Supplemental Figure 2B) expression of dual-FISH labeled cells. MUC5AC immunopositivity in Hi-VL subjects was ∼10-fold higher than in Lo-VL samples as quantified by mean fluorescence intensity (MFI) per cell (Figure 4G) whereas MUC5B expression did not significantly differ (Supplemental Figure 2C). Thus, both mucoinflammatory responses and immunomodulatory lncRNA expression of nasopharyngeal cells are significantly altered in Hi-VL SARS-CoV-2 infected patients as compared to Lo-VL patients. Current studies are ongoing to identify the molecular targets/binding partners of these lncRNAs.

**Figure 4.**
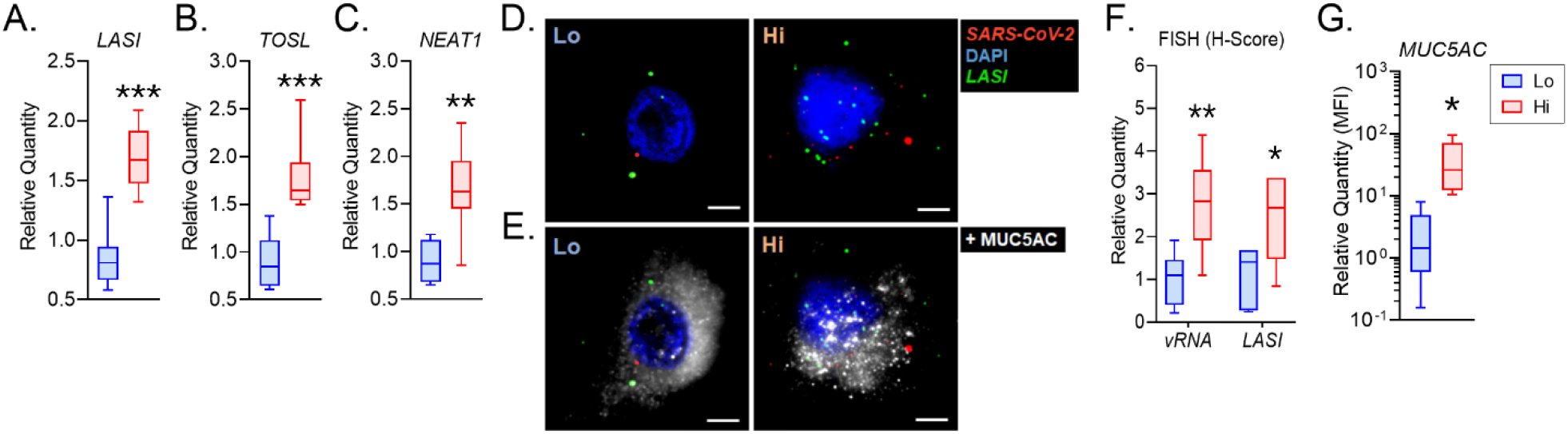
COVID-19 patients with high viral load show increased levels of immunomodulatory lncRNAs and the MUC5AC mucin expression. Relative expression of immunomodulatory lncRNAs, **(A.)** *LASI*, **(B.)** *TOSL*, and **(C.)** *NEAT1* in the Hi vs Lo patient swab samples. **(D.)** Colocalization of SARS-CoV-2 *vRNA* and *LASI* transcripts in nasal swab samples as detected by dual-FISH analysis. Representative micrographs from low (Lo) and high (Hi) viral load patient samples display detection of SARS-CoV-2 nucleocapsid *vRNA* (green) and *LASI* transcripts (red) along with DAPI-stained nuclei (blue). **(E.)** Nasal swab cells showing immunoreactive MUC5AC expression (shown in white) in the dual-FISH labeled cells, scale – 2 µ. **(F.)** H-score quantitation of *vRNA* and *LASI* transcripts in COVID-19 patients. **(G.)** Relative quantitation of mean fluorescence intensity (MFI) of MUC5AC expression in Lo-VL and Hi-VL patient samples. (n=10/gp for data in A, B, C, & F; and n=4/gp for data in G; *p<0.05; \*\**p*<0.01; \*\*\**p*<0.001 by Student’s t-test)

We next performed *in-silico* analysis of FASTQ files obtained from RNA-sequencing of lung biopsies from COVID-19 patients and healthy controls as earlier reported (32, 33). Notably, although only a limited number of samples were analyzed, there was a markedly higher expression of *MUC5AC, MUC5B*, and *SPDEF* genes in COVID-19 patients (Supplemental Figure 3A) whereas there were no changes in *IL-6, CXCL-8*, and *ICAM-1* expressions (Supplemental Figure 3B). In terms of lncRNAs, *MALAT1* trended lower in COVID-19 patients, *LASI* lncRNA expression trended higher and was significantly elevated in at least one COVID-19 patient, and there was no observed difference in *NEAT1* lncRNA levels between COVID-19 patients and healthy controls (Supplemental Figure 3C). That said, these data do further corroborate the association of mucoinflammatory response and lncRNAs with COVID-19 pathophysiology observed in our 3D models and clearly warrant further analyses of larger data sets and the underlying molecular mechanisms.

In conclusion, the present study employing both a 3D airway tissue model of SARS-CoV-2 infection and COVID-19 patient samples provides the first experimental association of SARS-CoV-2 viral load with mucoinflammatory phenotype and specific airway mucosa lncRNAs despite limited cohort sample size, possible variation in swab sampling, and/or the cell numbers per sample potentially contributing to data incongruity. Importantly, we have recently shown that blocking lncRNA *LASI* can help suppress mucus hyperexpression (34) and hence, could be a feasible strategy to regulate COVID-19 associated mucoinflammatory response. As initial host-viral interactions in the upper respiratory tract are crucial for modulating subsequent respiratory and systemic immune responses to SARS-CoV-2 and other co-infections and environmental exposures; targeting these lncRNAs could potentially rescue the debilitating outcomes of the COVID-19 sequelae and help improve current disease management strategies.

## Methods

For more details, see Supplemental Methods.

### Statistics

Statistical methods are described in Supplemental Methods. Briefly, grouped results were expressed as mean ± SEM and *P* values less than or equal to 0.05 were considered significant.

### Study approval

The COVID-19 patient samples were collected at University of Miami Biobank facility according to the approved institutional review board protocol. All studies involving SARS-CoV-2 wild type virus were conducted in the University of Nebraska Medical Center (UNMC) BSL-3 high containment core facility according to the institutional biosafety committee approved protocol. The formalin-fixed and frozen samples were analyzed at the Florida International University according to the approved institutional biosafety committee protocols in a designated BSL-2+ facility.

## Data Availability

The data that support the findings of this study are available on request from the corresponding author. The online datasets analyzed in this study are openly available as FASTQ files i.e. SRX8089341, SRX8089342, SRX8089343 and SRX8089344 files.

## Authors’ contributions

Conception and study design: HSC

Formalizing hypothesis: HSC, GMB, SNB

Patient samples and BSL3 experiments: MM2, AA, KP, SNB

Data acquisition: DD, AA, MM1, KP

Data analysis: DD, AA, MM1, KP

Data interpretation: DD, AA, MM1, KP, GMB, MN, MM2, SNB, HSC

Drafting the manuscript for important intellectual content: DD, HSC, SNB

Reviewing manuscript: DD, AA, MM1, KP, GMB, MN, MM2, SNB, HSC

Approving final version of manuscript for submission: HSC

## Acknowledgements

We thank the staff of the University of Miami Biobank facility and of the University of Nebraska Medical Center BSL-3 high containment core facility for helping us to carry out these studies.

## Abbreviations Used

ACE2: angiotensin-converting enzyme 2
ARDS: acute respiratory distress syndrome
COVID-19: coronavirus infection disease of 2019
C_T_: cycle threshold from qRT-PCR
CXCL-8: C-X-C chemokine ligand-8
FISH: fluorescent in-situ hybridization
FOXA3: Forkhead Box A3 transcription factor
Hi-VL: high viral load
hpi: hours post-infection
ICAM-1: intercellular adhesion molecule-1
IL-6: interleukin-6
lncRNA: long noncoding RNA
LASI: lncRNA on antisense strand to ICAM-1
Lo-VL: low viral load
NEAT1: Nuclear Enriched Abundant Transcript 1
MALAT1: Metastasis Associated Lung Adenocarcinoma Transcript 1
MUC2: mucin 2, oligomeric mucus/gel-forming mucin
MUC4: mucin 4, cell surface associated mucin
MUC5AC: mucin 5AC, oligomeric mucus/gel-forming mucin
MUC5B: mucin 5B, oligomeric mucus/gel-forming mucin
RTECs: respiratory tract epithelial cells
SARS-CoV-2: severe acute respiratory syndrome coronavirus 2
SCGB1A1: secretoglobulin 1A1
SPDEF: SAM Pointed Domain Containing ETS Transcription Factor
TMPRSS2: transmembrane serine protease 2
TOSL: TNFAIP3-opposite strand lncRNA
vRNA: viral RNA

## REFERENCES

1. Hoffmann M, Kleine-Weber H, Schroeder S, Kruger N, Herrler T, Erichsen S, et al. SARS-CoV-2 Cell Entry Depends on ACE2 and TMPRSS2 and Is Blocked by a Clinically Proven Protease Inhibitor. Cell. 2020;181(2):271–80 e8.

2. Hou YJ, Okuda K, Edwards CE, Martinez DR, Asakura T, Dinnon KH, 3rd, et al. SARS-CoV-2 Reverse Genetics Reveals a Variable Infection Gradient in the Respiratory Tract. Cell. 2020;182(2):429–46 e14.

3. Zou L, Ruan F, Huang M, Liang L, Huang H, Hong Z, et al. SARS-CoV-2 Viral Load in Upper Respiratory Specimens of Infected Patients. N Engl J Med. 2020;382(12):1177–9.

4. Sungnak W, Huang N, Becavin C, Berg M, Queen R, Litvinukova M, et al. SARS-CoV-2 entry factors are highly expressed in nasal epithelial cells together with innate immune genes. Nat Med. 2020;26(5):681–7.

5. Radzikowska U, Ding M, Tan G, Zhakparov D, Peng Y, Wawrzyniak P, et al. Distribution of ACE2, CD147, CD26, and other SARS-CoV-2 associated molecules in tissues and immune cells in health and in asthma, COPD, obesity, hypertension, and COVID-19 risk factors. Allergy. 2020;75(11):2829–45.

6. Matusiak M, and Schurch CM. Expression of SARS-CoV-2 entry receptors in the respiratory tract of healthy individuals, smokers and asthmatics. Respir Res. 2020;21(1):252.

7. Qiang XL, Xu P, Fang G, Liu WB, and Kou Z. Using the spike protein feature to predict infection risk and monitor the evolutionary dynamic of coronavirus. Infect Dis Poverty. 2020;9(1):33.

8. Lukassen S, Chua RL, Trefzer T, Kahn NC, Schneider MA, Muley T, et al. SARS-CoV-2 receptor ACE2 and TMPRSS2 are primarily expressed in bronchial transient secretory cells. EMBO J. 2020;39(10):e105114.

9. Schuler BA, Habermann AC, Plosa EJ, Taylor CJ, Jetter C, Negretti NM, et al. Age-determined expression of priming protease TMPRSS2 and localization of SARS-CoV-2 in lung epithelium. J Clin Invest. 2021;131(1).

10. Hewitt RJ, and Lloyd CM. Regulation of immune responses by the airway epithelial cell landscape. Nat Rev Immunol. 2021.

11. Bhatraju PK, Ghassemieh BJ, Nichols M, Kim R, Jerome KR, Nalla AK, et al. Covid-19 in Critically Ill Patients in the Seattle Region - Case Series. N Engl J Med. 2020;382(21):2012–22.

12. Devadoss D, Long C, Langley RJ, Manevski M, Nair M, Campos MA, et al. Long Noncoding Transcriptome in Chronic Obstructive Pulmonary Disease. Am J Respir Cell Mol Biol. 2019;61(6):678–88.

13. Yao RW, Wang Y, and Chen LL. Cellular functions of long noncoding RNAs. Nat Cell Biol. 2019;21(5):542–51.

14. Mukherjee S, Banerjee B, Karasik D, and Frenkel-Morgenstern M. mRNA-lncRNA Co-Expression Network Analysis Reveals the Role of lncRNAs in Immune Dysfunction during Severe SARS-CoV-2 Infection. Viruses. 2021;13(3).

15. Cheng J, Zhou X, Feng W, Jia M, Zhang X, An T, et al. Risk stratification by long non-coding RNAs profiling in COVID-19 patients. J Cell Mol Med. 2021;25(10):4753–64.

16. Morenikeji OB, Bernard K, Strutton E, Wallace M, and Thomas BN. Evolutionarily Conserved Long Non-coding RNA Regulates Gene Expression in Cytokine Storm During COVID-19. Front Bioeng Biotechnol. 2020;8:582953.

17. Meydan C, Madrer N, and Soreq H. The Neat Dance of COVID-19: NEAT1, DANCR, and Co-Modulated Cholinergic RNAs Link to Inflammation. Front Immunol. 2020;11:590870.

18. Blanco-Melo D, Nilsson-Payant BE, Liu WC, Uhl S, Hoagland D, Moller R, et al. Imbalanced Host Response to SARS-CoV-2 Drives Development of COVID-19. Cell. 2020;181(5):1036–45 e9.

19. Vishnubalaji R, Shaath H, and Alajez NM. Protein Coding and Long Noncoding RNA (lncRNA) Transcriptional Landscape in SARS-CoV-2 Infected Bronchial Epithelial Cells Highlight a Role for Interferon and Inflammatory Response. Genes (Basel). 2020;11(7).

20. Hao S, Ning K, Kuz CA, Vorhies K, Yan Z, and Qiu J. Long-Term Modeling of SARS-CoV-2 Infection of In Vitro Cultured Polarized Human Airway Epithelium. mBio. 2020;11(6).

21. Ridley C, and Thornton DJ. Mucins: the frontline defence of the lung. Biochem Soc Trans. 2018;46(5):1099–106.

22. Weitnauer M, Mijosek V, and Dalpke AH. Control of local immunity by airway epithelial cells. Mucosal Immunol. 2016;9(2):287–98.

23. Park SJ, Lee K, Kang MA, Kim TH, Jang HJ, Ryu HW, et al. Tilianin attenuates HDM-induced allergic asthma by suppressing Th2-immune responses via downregulation of IRF4 in dendritic cells. Phytomedicine. 2021;80:153392.

24. Ma J, Rubin BK, and Voynow JA. Mucins, Mucus, and Goblet Cells. Chest. 2018;154(1):169–76.

25. Kesimer M, Ford AA, Ceppe A, Radicioni G, Cao R, Davis CW, et al. Airway Mucin Concentration as a Marker of Chronic Bronchitis. N Engl J Med. 2017;377(10):911–22.

26. Evans CM, Raclawska DS, Ttofali F, Liptzin DR, Fletcher AA, Harper DN, et al. The polymeric mucin Muc5ac is required for allergic airway hyperreactivity. Nat Commun. 2015;6:6281.

27. Chatterjee M,van Putten JPM, and Strijbis K. Defensive Properties of Mucin Glycoproteins during Respiratory Infections-Relevance for SARS-CoV-2. mBio. 2020;11(6).

28. Bonser LR, and Erle DJ. Airway Mucus and Asthma: The Role of MUC5AC and MUC5B. J Clin Med. 2017;6(12).

29. Roy MG, Livraghi-Butrico A, Fletcher AA, McElwee MM, Evans SE, Boerner RM, et al. Muc5b is required for airway defence. Nature. 2014;505(7483):412–6.

30. Rajavelu P, Chen G, Xu Y, Kitzmiller JA, Korfhagen TR, and Whitsett JA. Airway epithelial SPDEF integrates goblet cell differentiation and pulmonary Th2 inflammation. J Clin Invest. 2015;125(5):2021–31.

31. Chen G, Korfhagen TR, Karp CL, Impey S, Xu Y, Randell SH, et al. Foxa3 induces goblet cell metaplasia and inhibits innate antiviral immunity. Am J Respir Crit Care Med. 2014;189(3):301–13.

32. Lu W, Liu X, Wang T, Liu F, Zhu A, Lin Y, et al. Elevated MUC1 and MUC5AC mucin protein levels in airway mucus of critical ill COVID-19 patients. J Med Virol. 2021;93(2):582–4.

33. Chu H, Hu B, Huang X, Chai Y, Zhou D, Wang Y, et al. Host and viral determinants for efficient SARS-CoV-2 infection of the human lung. Nat Commun. 2021;12(1):134.

34. Turjya RR, Khan MA, and Mir Md Khademul Islam AB. Perversely expressed long noncoding RNAs can alter host response and viral proliferation in SARS-CoV-2 infection. Future Virol. 2020;15(9):577–93.

35. Natarelli L, Parca L, Mazza T, Weber C, Virgili F, and Fratantonio D. MicroRNAs and Long Non-Coding RNAs as Potential Candidates to Target Specific Motifs of SARS-CoV-2. Noncoding RNA. 2021;7(1).

36. Devadoss D, Daly G, Manevski M, Houserova D, Hussain SS, Baumlin N, et al. A long noncoding RNA antisense to ICAM-1 is involved in allergic asthma associated hyperreactive response of airway epithelial cells. Mucosal Immunol. 2021;14(3):630–9.

37. Li G, Fan Y, Lai Y, Han T, Li Z, Zhou P, et al. Coronavirus infections and immune responses. J Med Virol. 2020;92(4):424–32.

38. Maltezou HC, Raftopoulos V, Vorou R, Papadima K, Mellou K, Spanakis N, et al. Association Between Upper Respiratory Tract Viral Load, Comorbidities, Disease Severity, and Outcome of Patients With SARS-CoV-2 Infection. J Infect Dis. 2021;223(7):1132–8.

39. Cui H, Banerjee S, Guo S, Xie N, Ge J, Jiang D, et al. Long noncoding RNA Malat1 regulates differential activation of macrophages and response to lung injury. JCI Insight. 2019;4(4).

40. Cai C, Qiu J, Qiu G, Chen Y, Song Z, Li J, et al. Long non-coding RNA MALAT1 protects preterm infants with bronchopulmonary dysplasia by inhibiting cell apoptosis. BMC Pulm Med. 2017;17(1):199.

41. Lee S, Kim T, Lee E, Lee C, Kim H, Rhee H, et al. Clinical Course and Molecular Viral Shedding Among Asymptomatic and Symptomatic Patients With SARS-CoV-2 Infection in a Community Treatment Center in the Republic of Korea. JAMA Intern Med. 2020;180(11):1447–52.

42. Fajnzylber J, Regan J, Coxen K, Corry H, Wong C, Rosenthal A, et al. SARS-CoV-2 viral load is associated with increased disease severity and mortality. Nat Commun. 2020;11(1):5493.

