## Supplemental Data for "Distinct Mucoinflammatory Phenotype and the Immunomodulatory Long Noncoding Transcripts Associated with SARS-CoV-2 Airway Infection"

**Address for Correspondence:**

Hitendra S. Chand, PhD

### METHODS

#### Production and Titration of SARS-CoV-2 Clinical Isolate

All studies involving the SARS-CoV-2 wild-type virus were conducted in the University of Nebraska Medical Center (UNMC) BSL-3 high containment core facility. SARS-CoV-2 isolate USA-WI1/2020 (BEI; cat# NR-52384) was passaged in Vero-STA-1 knockout cells. The viral titer was determined using the plaque assay (1). Briefly, Vero E6 cells were seeded in 6-well plates. After 24 h, cells were washed with sterile PBS (pH 7.4). The viral stock was serially diluted and added to cells in duplicate with new media and the plates were incubated at 37˚C for 1 h with occasional shaking every 15 m. Then 2 mL of 0.5% agarose in minimal essential media (MEM, Gibco, Thermofisher Inc) containing 5% FBS and antibiotics was added per well. Plates were incubated at 37°C humidified $CO_2$ incubator for 72 h. Then the cells were fixed with 4% paraformaldehyde (PFA, Pierce, ThermoFisher Scientific) overnight, followed by removing the overlay and then stained with 0.2% crystal violet to visualize plaque-forming units (PFU).

#### SARS-CoV-2 Infection of Airway Epithelial Cells

Primary human respiratory epithelial cells were either purchased from the Marsico Lung Institute Tissue Core (University of North Carolina, Chapel Hill) or MatTek Incorp (EpiAirway™, Ashland, MA). Cells were maintained in a bronchial epithelial growth medium (BEGM, Lonza, Walkersville, MD). For air-liquid interface (ALI) cultures, cells were plated onto collagen IV-coated 24 mm Transwell-clear culture inserts (Corning Costar Corporation, Cambridge, MA) at a density of 5 x $10^5$ cells/cm$^2$ in ALI media. The apical surface of transwells with cells was exposed to air and cells were cultured for another 21 d till they were fully differentiated. All the methods were performed in accordance with the institutional guidelines and regulations.

The 3-D airway tissue was infected with SARS-CoV-2 primary isolate (USA-WI1/2020, BEI Cat # NR-52384). Briefly, cells were infected with 1 MOI of SARS-CoV-2 inoculum prepared in ALI culture media from both apical and basal surface. The infected transwells were incubated for

1 h at 37°C in 5% $CO_2$ incubator and the plates were gently mixed every 15 m to facilitate uniform virus adsorption on to the cells. After 30 m the apical inoculum was removed and added to the basal media and after additional 30 m the total virus Inoculum was removed. The transwells were washed thoroughly with PBS thrice on both apical and basal surfaces and fresh media was added basally. Apical washes and the basal culture supernatants were collected at 1, 4, 24, and 48 h post-infection (hpi) and stored at -80°C for downstream analysis. For apical washes, 200 μl of PBS with 5 mM 2-mercaptoethanol (2-ME, Sigma Co.) was added to the apical surface of the cells and the apical wash was collected and cells were harvested after 30 m incubation. The aliquots of transwells were lyzed in either RIPA buffer with protease inhibitors (ThermoFisher Inc) or in RLT buffer (RNeasy kit, Qiagen Inc) with 143 mM 2-ME for total RNA isolation or were fixed with 4% PFA, washed with PBS and stored at 4°C for downstream application.

**Immunostaining and Fluorescent Imaging Analysis**

The formalin-fixed cells grown on transwells or the coated coverslips and infected with SARS-CoV-2 clinical isolate were fixed in 4% PFA and washed in 0.05% v Brij-35 in PBS (pH 7.4) and immunostaining was performed as described previously (2). Briefly, the cells were blocked using a solution containing 3% BSA, 1% Gelatin and 1% normal donkey serum with 0.1% Triton X-100 and 0.1% Saponin. They were stained with antibodies to MUC5AC (Millipore Inc., Burlington, MA), and MUC5B (Cell Signaling Tech., Danvers, MA) or isotype controls. The immunolabelled cells were detected using respective secondary antibodies conjugated fluorescent dyes (Jackson ImmunoResearch Lab Inc., West Grove, PA) and mounted with 4',6-diamidino-2-phenylindole (DAPI) containing Fluormount-G™ (SouthernBiotech, Birmingham, AL) for nuclear staining. Immunofluorescent images were captured using the BZX700 Microscopy system (Keyence Corp., Japan) and analyzed using NIH Image J software. The formalin-fixed nasal swab cells were also processed similarly following the washing with 0.05% v Brij-35 in PBS (pH 7.4) and were immunostained for MUC5AC and MUC5B. The expression was quantified by measuring the mean fluorescent intensity (MFI) per cell or the percent positive cells per treatment group.

**Quantitative Real-Time PCR for mRNAs and lncRNAs**

Total RNA was isolated from the snap-frozen tissue or cells using RNeasy kit (Qiagen, Germantown, MD) as per the manufacturer's instruction. RNA concentration was determined using the Synergy LX Multi-Mode reader (BioTek, Winooski, VT) and cDNA weas synthesized using iScript advanced cDNA kit (BioRad, Hercules, CA). The primer/probe sets for *MUC5AC, MUC5B, SCGB1A1, SPDEF, FOXA3, LASI, TOSL, NEAT1* and *MALAT1* transcripts were obtained from Applied Biosystems (Thermo Fisher Inc.) and cDNA amplified was quantified by q-PCR using the TaqMan Gene expression kit (Thermo Fisher Inc.). The SYBR green-based primer sets for *SARS-CoV-2 N1 viral RNA, ACE2, TMPRSS2, SCGB1A1, MUC2, MUC4, MUC5AC, MUC5B, NEAT1, ICAM-1, IL-6,* and *CXCL-8* mRNA were obtained from BioRad (Hercules, CA) and cDNA amplified by qPCR using the iTaq SYBR-green Master Mix (BioRad, Hercules, CA) in the BioRad CFX96 Real-Time PCR System (Hercules, CA). Relative quantities were calculated by normalizing averaged CT values to *U6* or *GAPDH* to obtain ΔCT, and the fold-change (ΔΔCT) over the controls were determined as described previously (3).

**Dual RNA Fluorescent In-Situ Hybridization (FISH)**

The dual-RNA FISH for LASI and SARS-CoV-2 N1 transcripts was essentially performed using the RNAscope® Fluorescent Multiplexed reagent kit (Advanced Cell Diagnostics, Newark, CA) as per the manufacturer's protocol. The custom-made probe sets for LASI and SARS-CoV-2 N1 were purchased from Advanced Cell Diagnostics (Newark, CA). Briefly, the formalin-fixed cells were permeabilized using 0.1% PBS Triton-X100 for 10 min at RT followed by washing in PBS and probed for the LASI transcript as described recently (2). Probes were hybridized for 2 h at 40$^{o}$C using a HyB-II EZ® oven. The signal was amplified using Tyramide signal amplification (TSA) reaction using TSA Plus Cy5 or Cy3 kit (PerkinElmer Bioscience) for *LASI* and *SARS-CoV-*

*2 N1 vRNA*, respectively, based on the manufacturer's instructions. The sections were either processed for immunostaining as described above or were directly stained with DAPI to visualize nuclei. Multifluorescent images were captured using a structured illumination module of the BZX700 Microscopy system (Keyence Corp, Japan) and analyzed by NIH ImageJ software.

RNA FISH expression was quantified by the analysis and as reported recently (3). Briefly, signals (dots/cell) for each transcript probe were counted and allocated to separate bins with Bin 0 (0 Dots/Cell); Bin 1 (1-3 Dots/Cell); Bin 2 (4-9 Dots/Cell); Bin 3 (10-15 Dots/Cell); Bin 4 (>15 Dots/Cell). The histology score (H-Score) was calculated as Sum of each (bin number x percentage of cells per bin) that ranged from 0 to 400 based on the transcript's expression, as described before (2).

**Enzyme-linked Immunosorbent Assays**

The interleukin (IL)-6 and the soluble ICAM-1 levels in culture supernatants were determined using a specific sandwich ELISAs kits as per the manufacturer's instructions. Briefly, the culture supernatants from SARS-CoV-2 infected cells were collected and centrifuged for measuring IL-6 (#430507, BioLegend Inc., San Diego, CA) and ICAM-1 (#LS-F4019, LifeSpan Biosciences Inc., Seattle, WA) in separate ELISAs. The absorbance was measured at 450 nm using a microplate reader (Synergy LX Multiplate Reader, BioTek Inc.) and the levels were calculated based on the standard curves.

**Dataset and Bioinformatics**

Raw RNA sequencing datasets were retrieved from the sequence read archive (SRA) database under accession no. PRJNA615032 (4). Detailed experimental procedures are explained in the aforementioned reference. Single-end FASTQ files for lung biopsies from healthy (SRX8089341 and SRX8089342) and from patients with COVID-19 (SRX8089343 and SRX8089344) were retrieved using the SRA toolkit version 2.9.2 as previously described (4, 5). FASTQ files (including both protein coding and non-coding RNAs) were aligned to the hg38 reference genome, transcript per million calculated (TPM) for each gene and heat maps generated as described previously (2).

**Statistical Analysis**

Grouped results were expressed as means ± SEM. Data were analyzed using GraphPad Prism v9.0 (GraphPad Software Inc., San Diego, CA). Grouped results were analyzed using a one-way analysis of variance and when significant main effects were detected ($p<0.05$), Fisher's least significant difference test was used to determine differences between groups. Student's t-test was used for data analysis between two groups.

### REFERENCES


1. Mendoza EJ, Manguiat K, Wood H, and Drebot M. Two Detailed Plaque Assay Protocols for the Quantification of Infectious SARS-CoV-2. *Curr Protoc Microbiol.* 2020;57(1):ecpmc105.

2. Devadoss D, Daly G, Manevski M, Houserova D, Hussain SS, Baumlin N, et al. A long noncoding RNA antisense to ICAM-1 is involved in allergic asthma associated hyperreactive response of airway epithelial cells. *Mucosal Immunol.* 2021;14(3):630-9.

3. Devadoss D, Singh SP, Acharya A, Do KC, Periyasamy P, Manevski M, et al. HIV-1 Productively Infects and Integrates in Bronchial Epithelial Cells. *Front Cell Infect Microbiol.* 2021.

4. Blanco-Melo D, Nilsson-Payant BE, Liu WC, Uhl S, Hoagland D, Moller R, et al. Imbalanced Host Response to SARS-CoV-2 Drives Development of COVID-19. *Cell.* 2020;181(5):1036-45 e9.

5. Vishnubalaji R, Shaath H, and Alajez NM. Protein Coding and Long Noncoding RNA (lncRNA) Transcriptional Landscape in SARS-CoV-2 Infected Bronchial Epithelial Cells Highlight a Role for Interferon and Inflammatory Response. *Genes (Basel).* 2020;11(7).

**Supplemental Table 1.** Demographics of COVID-19 patients whose nasopharyngeal swab samples were analyzed. Medical history was available for twelve patients.

| | Lo-VL | Hi-VL | *P-value* |
|---|---|---|---|
| **Subjects** | 10 (7) | 10 (5) | |
| **Sex (M/F)** | 6M/4F | 7M/3F | |
| **Age (Y)** | 61.5±4.0 | 63.1±3.8 | *0.8163* |
| **SARS-CoV-2 N1 vRNA ($C_T$)** | 35.5±0.7 | 25.9±1.3 | *<0.0001* |
| **Hospitalization** | 7 | 5 | |
| **Hypertension** | 5 | 5 | |
| **Diabetes** | 4 | 0 | |
| **Infectious Disease** | 1 | 2 | |
| **Pneumonia** | 5 | 5 | |
| **Oxygen Supplementation (>6L $O_2$)** | 1 | 5 | |
| **ICU** | 3 | 5 | |
| **ARDS** | 3 | 5 | |
| **Obesity** | 5 | 3 | |

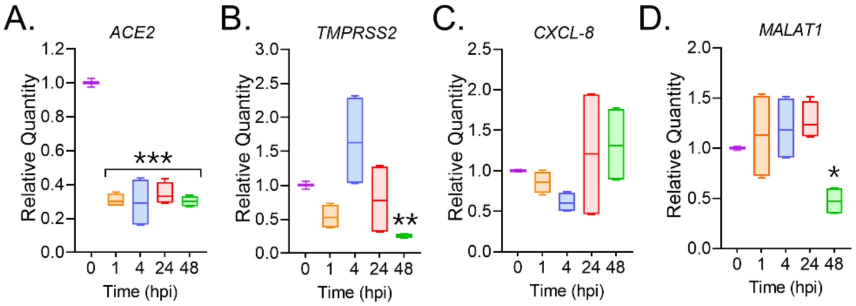


**<u>Supplemental Figure 1.</u> SARS-CoV-2 infection of human respiratory epithelial cells suppress ACE2 expression with no changes in TMPRSS2, CXCL-8 and MALAT1 lncRNA expression.** Relative expression levels of host viral entry regulating factors, **(A.)** ACE1 and **(B.)** TMPRSS2, and **(C.)** the inflammatory factors *CXCL-8* mRNA and **(D.)** the *MALAT-1 lncRNA* expression analyzed at 0, 1, 4, 24, and 48 h post-infection (hpi) with SARS-CoV-2 clinical isolate. (n=4/gp from 2 independent experiments; *p<0.05; ***p*<0.01;****p*<0.001 by ANOVA)

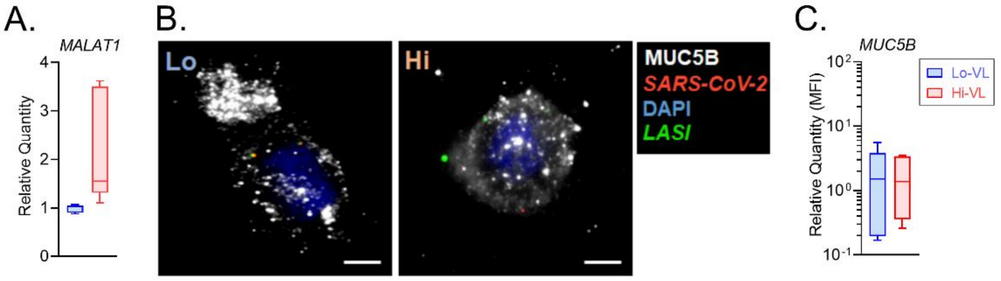


**<u>Supplemental Figure 2.</u> The intracellular MUC5B mucin expression levels are similar in Hi vs Lo viral load COVID-19 nasopharyngeal cells. (A.)** Relative expression of *MALAT-1* lncRNA in nasopharyngeal swab samples of COVID-19 patients. **(B.)** Representative micrographs showing immunoreactive MUC5B expression (shown in white) in the dual-FISH labeled cells displaying detection of SARS-CoV-2 nucleocapsid *vRNA* (green) and *LASI* transcripts (red) along with DAPI-stained nuclei (blue), scale – 2 µ. **(C.)** Relative quantitation of MUC5B expression (MFI) in low (Lo-VL) and high (Hi-VL) viral load patient samples (n=4/gp).

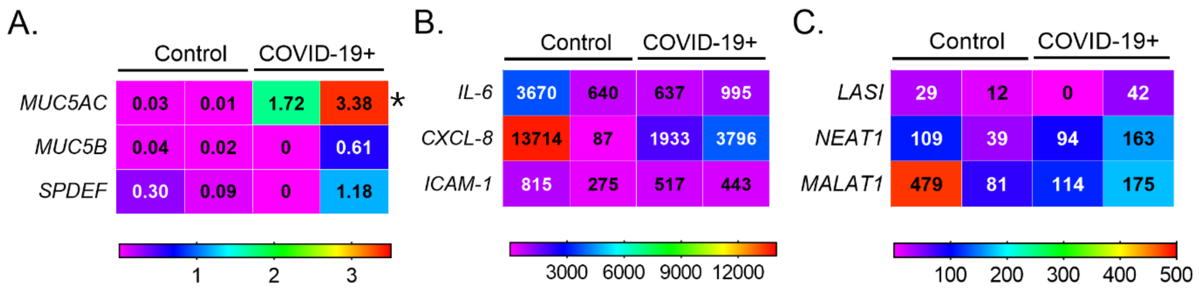


**<u>Supplemental Figure 3.</u> Relative expression of respiratory mucins, inflammatory factors, and select lncRNAs in COVID-19 patients and healthy control lung tissue.** Each column represents one patient with relative expression depicted in terms of the corresponding color scale and in transcripts per million (TPM) indicated for each gene (row). **(A.)** Expression of *MUC5AC, MUC5B, and SPDEF* mRNAs. **(B.)** Expression of *IL-6, CXCL-8, and ICAM-1 m*RNAs. **(C.)** Expression of *LASI, NEAT1, and MALAT1 lnc*RNAs.
